# Performance evaluation of new fluorescent-based lateral flow immunoassay for quantification of HbA1c in diabetic patients

**DOI:** 10.1101/2022.10.27.22281596

**Authors:** Nadin Younes, Mahmoud M. Al Ghwairi, Amin F. Majdalawieh, Nader Al-Dweik, Gheyath K. Nasrallah

## Abstract

**Background:** Rapid and constant HbA1c level monitoring is essential in slowing the progression of Type 2 diabetes. This need becomes challenging in low resources countries where the social burden of the disease is overwhelming. Recently, fluorescent-based lateral flow immunoassays (LFIAs) gained wide attention for small or non-laboratory settings and population surveillance.

**Aim:** This study aim to evaluate the performance of novel fluorescence-based LFIA Finecare™ HbA1c Rapid Quantitative Test for quantitative measurement of HbA1c along with its reader (Model No. FS-113).

**Methods:** We conducted a retrospective study using 147 samples (fingerstick and venepuncture whole blood samples) analysed by Finecare™ HbA1c Rapid Quantitative Test. For validating Finecare™ measurements, results were compared with results of the reference assay: Roche Cobas Pro c 503.

**Results:** Finecare™ showed 92.7% sensitivity and 94.7% specificity compared to the Roche Cobas Pro c 503 using fingerstick whole blood samples. On the other hand, Finecare™ showed 98.7% sensitivity and 100% specificity compared to the Roche Cobas Pro c 503 using venepuncture blood samples. Cohen’s Kappa statistic denoted excellent agreement with Roche Cobas Pro c 503, with values being 0.84 (95% CI: 0.72-0.97) and 0.97 (95% CI: 0.92-1.00) using fingerstick whole blood samples and venous blood, respectively. In addition, a strong correlation was observed between Finecare™/Roche Cobas Pro c 503 (*r>*0.9, *p*<0.0001) with fingerstick and venous blood samples. Most importantly, Finecare™ showed a significant difference between the normal, pre-diabetic, and diabetic samples (*p*<0.001).

**Conclusion:** Finecare™ is a reliable assay and can be easily implemented for long-term monitoring of HbA1c in diabetic patients, particularly in none or small laboratory settings.

## 1. Introduction

Glycated haemoglobin (HbA1c) serves as a reliable indicator of glycemic status in diabetic patients over a period of two to three months (1). HbA1c is produced once haemoglobin is chemically linked to glucose (1). Traditionally, high plasma glucose levels were used for diagnosis of diabetes mellitus. This could be done after fasting, two hours after an oral glucose (75 g) tolerance test, or after a random blood glucose check in symptomatic patients (2). Recently, the American Diabetes Association and the World Health Organisation (WHO) recommended the use of HbA1c (≥ 6.5%) for the diagnosis of diabetes mellitus (3). This was based on the fact that HbA1c’s can predict clinical outcomes of the disease. In this context, many studies showed that HbA1c has a strong correlation with the chronic microvascular complications of diabetes, including retinopathy, nephropathy and neuropathy (4, 5).

Despite the fact that HbA1c testing is currently more expensive than blood glucose testing (the average net cost of a HbA1c test is 13.6 times that of a plasma glucose measurement), it offers substantial practical benefits (6). Most importantly, HbA1c testing can be conducted at any time throughout the day and does not require any special pre-test preparation by the patient (such as overnight fasting) (6, 7). Therefore, rapid and constant monitoring HbA1c helps in slowing the progression of type-2 diabetes in patients. However, this need becomes a challenge in limited-resource settings lacking laboratory infrastructure, where the social burden of the disease is often overpowering (8). The current laboratory diagnostic techniques for HbA1c such as cation-exchange HPLC, affinity chromatography, and capillary electrophoresis, involve expensive instruments, laborious and require a long turnaround time (9). Under these conditions, constant testing and monitoring can be facilitated by point-of-care (POC) devices. However, existing POC devices for HbA1c often require trained personnel and large equipment and are therefore not suited for non-laboratory testing.

Lateral flow immunoassays (LFIAs) are attractive for small or point of care settings and population surveillance. They are rapid, in expensive, simple to use, most importantly, rely on easily accessible samples such as whole blood from fingerstick. The Finecare™ HbA1c Rapid Quantitative Test is a fluorescence immunoassay used along with Finecare™ FIA System for quantitative determination of HbA1c in human blood (venepuncture or fingerstick). In this study, we aimed to evaluate the performance of Finecare™ HbA1c Rapid Quantitative Test by using samples obtained by fingerstick and venepuncture. In addition, to compare the performance of Finecare™ HbA1c Rapid Quantitative Test with the reference technique; Roche Cobas pro c 503 clinical chemistry analyzer from Roche Diagnostics.

## 2. Methods

### 2.1 Sample collection and ethical approval

This is a retrospective study. Thus, no patients/applicants were recruited and there was no direct or indirect interaction with any human subject. The study was conducted on existing de-identified testing results obtained from the manufacturer to conduct the analysis. Fingerstick and matched venous blood sample was drawn from a total of 147 participants in two different laboratories (one private lab and another lab that belongs to the ministry of public health in Jordan). Ethical approval exemption has been granted (QU-IRB 1766-E/22) from Qatar University.

### 2.2 Finecare™ HbA1c rapid quantitative test

The Finecare™ HbA1c Rapid Quantitative Test is based on fluorescence immunoassay technology. The test uses a sandwich immunodetection method to measure percentage of HbA1c in human blood. Briefly, 20 μL of fingerstick or venepuncture blood was added into detection buffer tube. After that, 75 μL of sample mixture was loaded into the sample well and was inserted into the test cartridge holder of Finecare™ FIA meters. The reaction time is 5 minutes.

### 2.3 Reference method Roche Cobas Pro c 503

The Tina-quant Hemoglobin A1cDx assay is intended to diagnose diabetic patients. It is in vitro diagnostics assay to quantify hemoglobin A1c (mmol/mol) and % hemoglobin A1c in hemolysate or venous whole blood on the cobas c 503 clinical chemistry analyzers. This approach is based on the turbidimetric inhibition immunoassay of blood samples that have been hemolyzed. The anti-HbA1c antibody forms a soluble complex with a single binding site on HbA1c. Polyhaptens react with excess anti-HbA1c antibody to generate an insoluble compound, which is evaluated by turbidimetry.

### 2.5 Statistical method

Correlation and linear regression analysis were conducted between Finecare™ and the reference method; Roche Cobas Pro c 503. Because our data was not normally distributed, spearman correlation coefficient (r) was calculated. For absolute values of spearman’s r, 0– 0.19 is denoted as very weak, 0.2–0.39 as weak, 0.40–0.59 as moderate, 0.6–0.79 as strong, and 0.8–1 as very strong correlation (10).

Concordance analysis between Finecare™ and the reference methods Roche Cobas Pro c 503 was conducted, which includes the overall percent agreement (OPA), positive percent agreement (PPA), and negative percent agreement (NPA), accuracy/efficiency as well as Cohen’s Kappa statistic, which is a robust metric that estimates the level of agreement between two diagnostic tests. A Cohen’s Kappa coefficient <0.40 suggests a poor agreement, 0.40–0.59 suggests a fair agreement, 0.60–0.74 suggests a good agreement, and ≥0.75 suggests an excellent agreement (11). Finally, we assessed the area under (a ROC) curve, which measures the accuracy of a quantitative diagnostic test (12). An AUC of 0.9–1.0 is denoted as excellent, 0.8–0.9 is denotes as very good, 0.7–0.8 is denoted as good, 0.6–0.7 is denoted as sufficient, 0.5–0.6 is denoted as bad, and <0.5 is denoted as not useful. The significance level was indicated at 5%, and a 95% confidence interval (CI) was reported for each metric. All statistical analysis was performed using GraphPad Prism software (Version 9, San Diego, CA, USA).

## 3. Results

### 3.1 Finecare™ results are comparable to the standard laboratory method Roche Cobas Pro c 503

We assessed the performance of Finecare™ using fingerstick and venepuncture whole blood samples in comparison to Roche Cobas Pro c 503.

The overall distribution of the values generated by each automated analyzer against the cut-offs (dashed lines) is shown in **Figure 1**. As depicted in **Figure 1**, there was no significant difference between results obtained from Finecare™ and Roche Cobas Pro c 503 analytical analyser using the fingerstick or venous blood samples.

**Figure 1.**
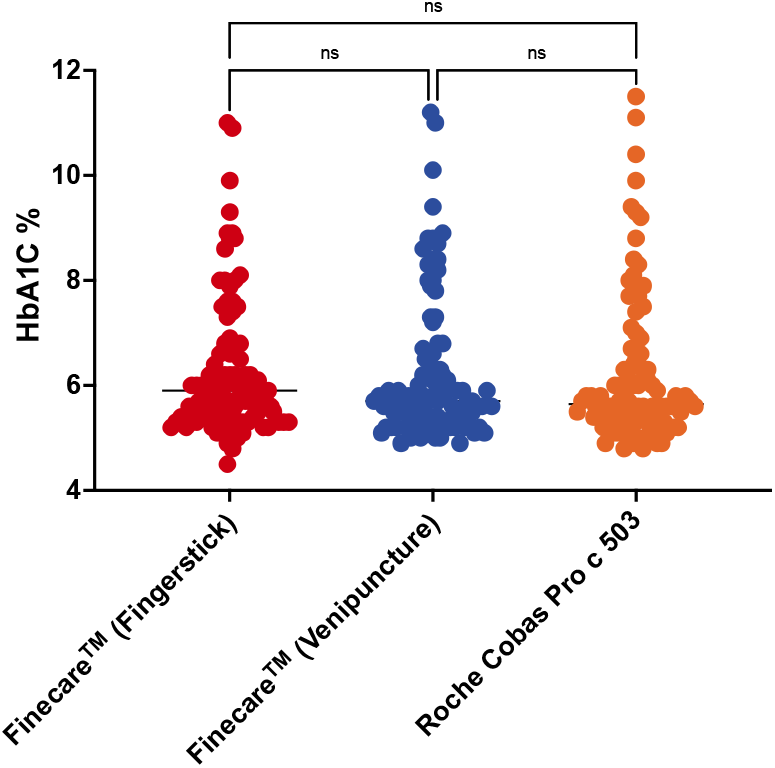
Distribution of numerical results obtained from fingerstick whole blood and venous blood using Finecare™ machine and the reference method; Roche Cobas Pro c 503. Nonparametric Kruskal-Wallis test was used to compare the differences between groups.

### 3.2 Finecare™ showed low level of false positive and false negative comparable to that of the standard laboratory method

According to the current recommendations, patients with a HbA1c level ≥6.5% would get intensive treatment. To assess the clinical application of Finecare™, the HbA1c results were compared with those of the conventional laboratory results using a cut-off point of 6.5% for HbA1c using fingerstick and venous blood samples, as shown in Table 1 and Table 2. Using fingerstick whole blood, 4 samples showed false positive results (4%) and two samples (2%) showed false negative results using Finecare™ as shown in Table 1. Whereas, using venous blood samples, only one sample showed false positive result (1%) and none of the samples showed false negative results (0%) using Finecare™ as shown in Table 2. Similarly, as shown in Table 3, only 3 fingerstick whole blood samples showed false positive results (3%) when compared to venous blood, whereas only 2 fingerstick whole blood samples showed false negative results (2%) when compared to venous blood samples.

**Table 1.** Comparison between Roche Cobas Pro c 503 and Finecare™ (fingerstick) tests.

| Cut-off: 6.5% |  | Reference Method<br>Roche Cobas Pro c 503 |  |  |
| --- | --- | --- | --- | --- |
|  |  | POSITIVE | NEGATIVE | TOTAL |
| **Finecare<sup>TM</sup>**<br>**(fingerstick)** | POSITIVE | 23 | 4 | 27 |
|  | NEGATIVE | 2 | 71 | 73 |
|  | TOTAL | 25 | 75 | 100 |

**Table 2.** Comparison between Roche Cobas Pro c 503 and Finecare™ (venepuncture) tests.

| Cut-off: 6.5% |  | Reference Method<br>Roche Cobas Pro c 503 |  |  |
| --- | --- | --- | --- | --- |
|  |  | POSITIVE | NEGATIVE | TOTAL |
| **Finecare<sup>TM</sup>**<br>**(venepuncture)** | POSITIVE | 25 | 1 | 26 |
|  | NEGATIVE | 0 | 74 | 74 |
|  | TOTAL | 25 | 75 | 100 |

**Table 3.** Comparison between Finecare™ (venepuncture) and Finecare™ (fingerstick) tests.

| Cut-off: 6.5% |  | Finecare<sup>TM</sup> (venepuncture) |  |  |
| --- | --- | --- | --- | --- |
|  |  | POSITIVE | NEGATIVE | TOTAL |
| **Finecare<sup>TM</sup>** | POSITIVE | 24 | 3 | 27 |
| (fingerstick) | NEGATIVE | 2 | 71 | 73 |
|  | TOTAL | 26 | 74 | 100 |

### 3.3 Finecare™ demonstrates high sensitivity and specificity comparable to that of the standard laboratory method

As shown in Table 4, Finecare™ demonstrated 100% sensitivity and 98.7% specificity compared to the standard laboratory method using venous blood samples. Whereas Finecare™ showed lower sensitivity (92.0%) and specificity (94.7%) compared to the reference method using fingerstick whole blood. In addition, we evaluated the sensitivity and specificity of Finecare™ using fingerstick in comparison to venous blood samples. As expected, using fingerstick whole blood showed lower sensitivity (92.3%) and specificity (95.9%) compared to venous blood.

**Table 4.**
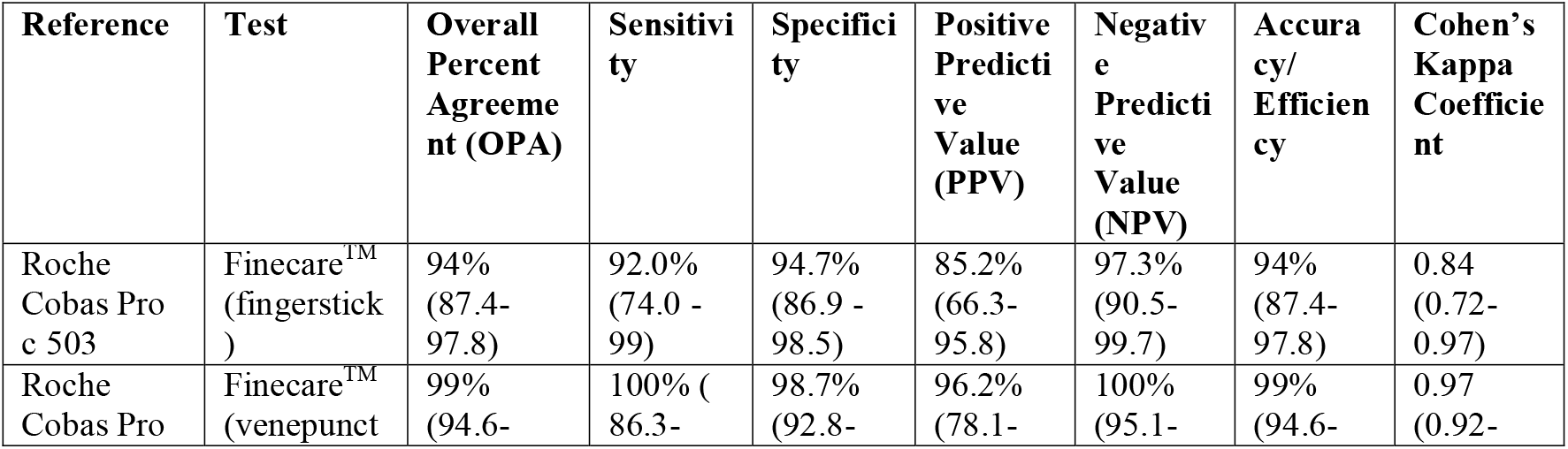
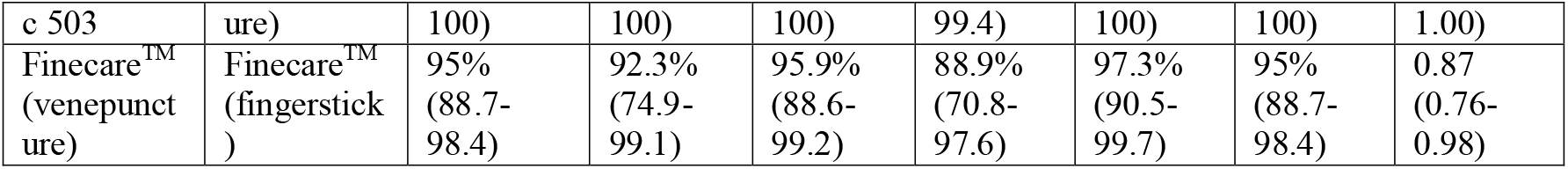
Concordance assessment between Finecare™ fingerstick, venepuncture test and Roche Cobas Pro c 503.

The concordance assessment between the reference method Roche Cobas Pro c 503 and Finecare™ using venous and fingerstick whole blood samples is reported in Table 4. The tests’ agreements were studied in a pairwise fashion applying inter-rater agreement statistics; (Cohen’s Kappa coefficient, κ). The OPA, PPV, and NPV between Roche Cobas Pro c 503 and Finecare™ using venous blood were 99%, 98.67%, and 100%, respectively, whereas using fingerstick whole blood were 94%, 94.67%, and 92%, respectively. Most importantly, Cohen’s Kappa coefficient denoted excellent agreement between Finecare™ and Roche Cobas Pro c 503 (κ >0.75) using fingerstick and venous whole blood samples.

### 3.4 Very strong correlation between Finecare™ and the standard laboratory method Roche Cobas Pro c 503

We assessed the correlation between Finecare™ with the standard laboratory method; Roche Cobas Pro c 503 using fingerstick and venous blood samples as shown in **Figure 2**. A very strong correlation was observed (*r*=0.95, *p*<0.0001) between Finecare™ using fingerstick whole blood sample and Roche Cobas Pro c 503. Similarly, very strong correlation was observed (*r*=0.97, *p*<0.0001) between Finecare™ using venous whole blood sample and Roche Cobas Pro c 503. The linear regression analysis showed that constructed model could strongly predict the dependent variable (R^2^=0.9, *p*<0.0001) between Finecare™ using venous or fingerstick samples.

**Figure 2.**
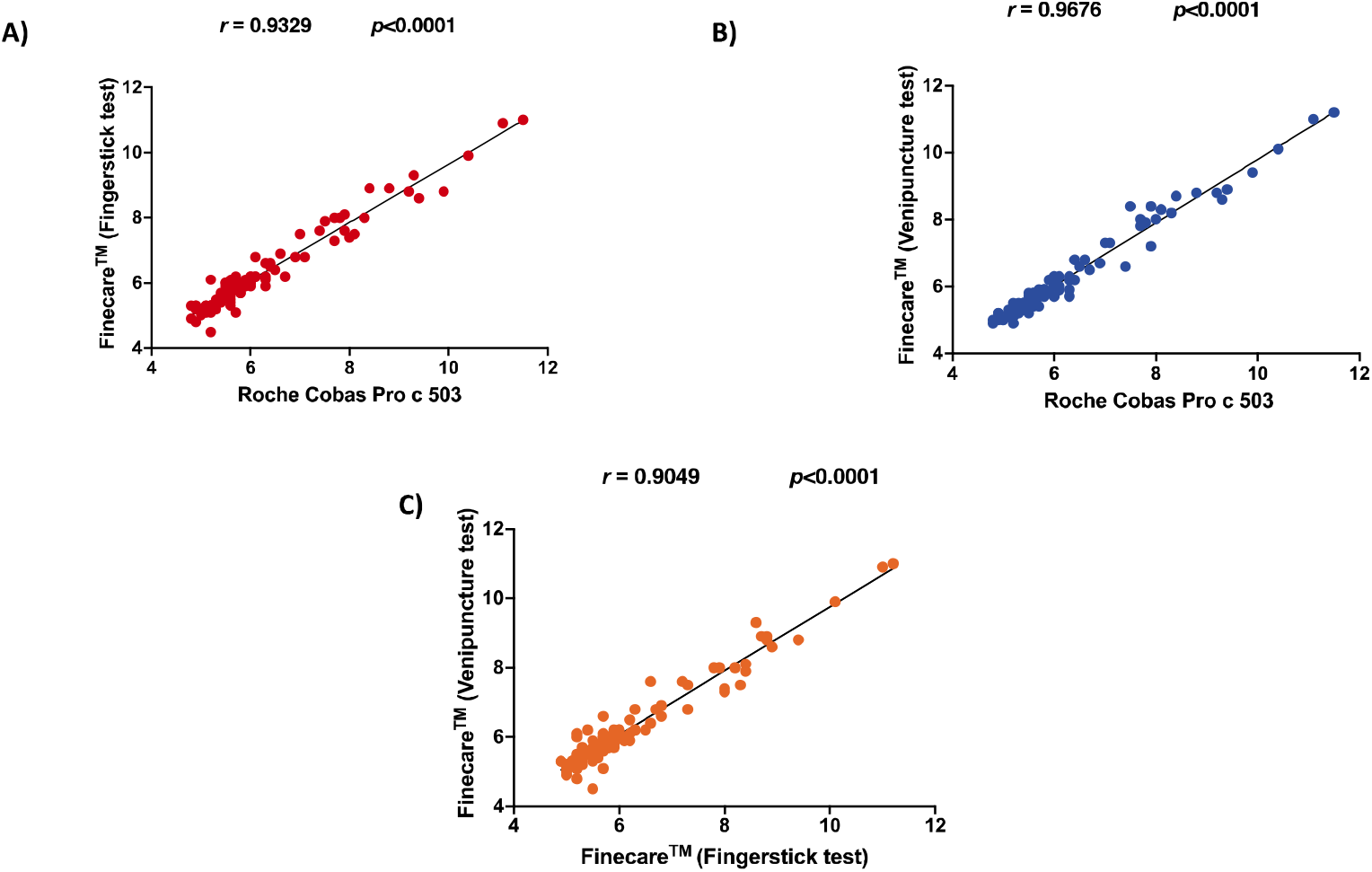
Pairwise correlation analysis and linear regression analysis of the numerical values obtained by each assay. Spearman correlation coefficient (r), and p value are indicated. Coefficient of determination (R^2^) was calculated to be 0.952, 0.969, 0.937 for fig A, B, and C, respectively.

Receiver-Operating Characteristic (ROC) curve analyses showed excellent performance for Finecare™ with an Area Under the Curve (AUC) of 0.994 and 0.998 using fingerstick and venous blood samples, respectively (**Figure 3**).

**Figure 3.**
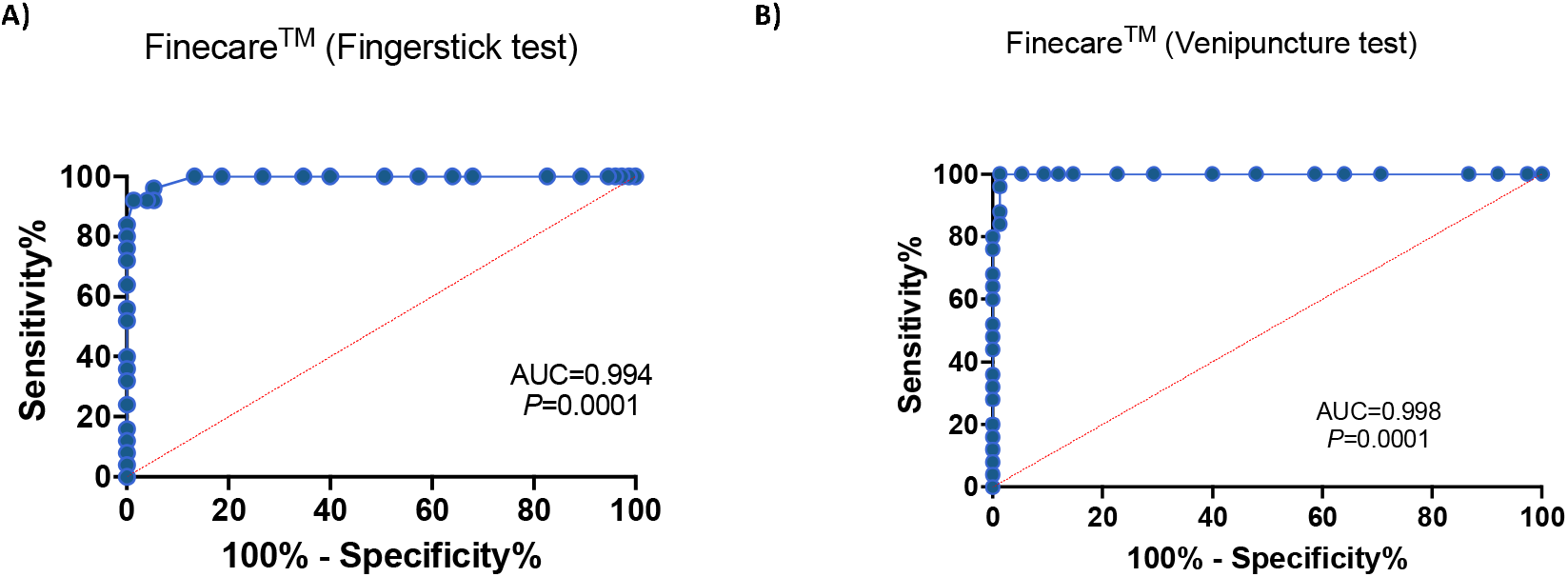
ROC curve for Finecare™. An AUC of 0.9–1.0 is considered excellent. AUC, area under the curve; ROC, Receiver Operating Characteristic.

### 3.5 Finecare™ test could distinguish between the pre-diabetic and diabetic groups similar to the standard laboratory method

We classified the participants into three groups according to ADA (American Diabetes Association): <5.7% (no diabetes), 5.7-6.5% (pre-diabetes), ≥6.5% (diabetes). Similar to the reference method, Finecare™ showed a significant difference between the normal, pre-diabetic, and diabetic samples (*p*<0.001) as shown in **Figure 4**.

**Figure 4.**
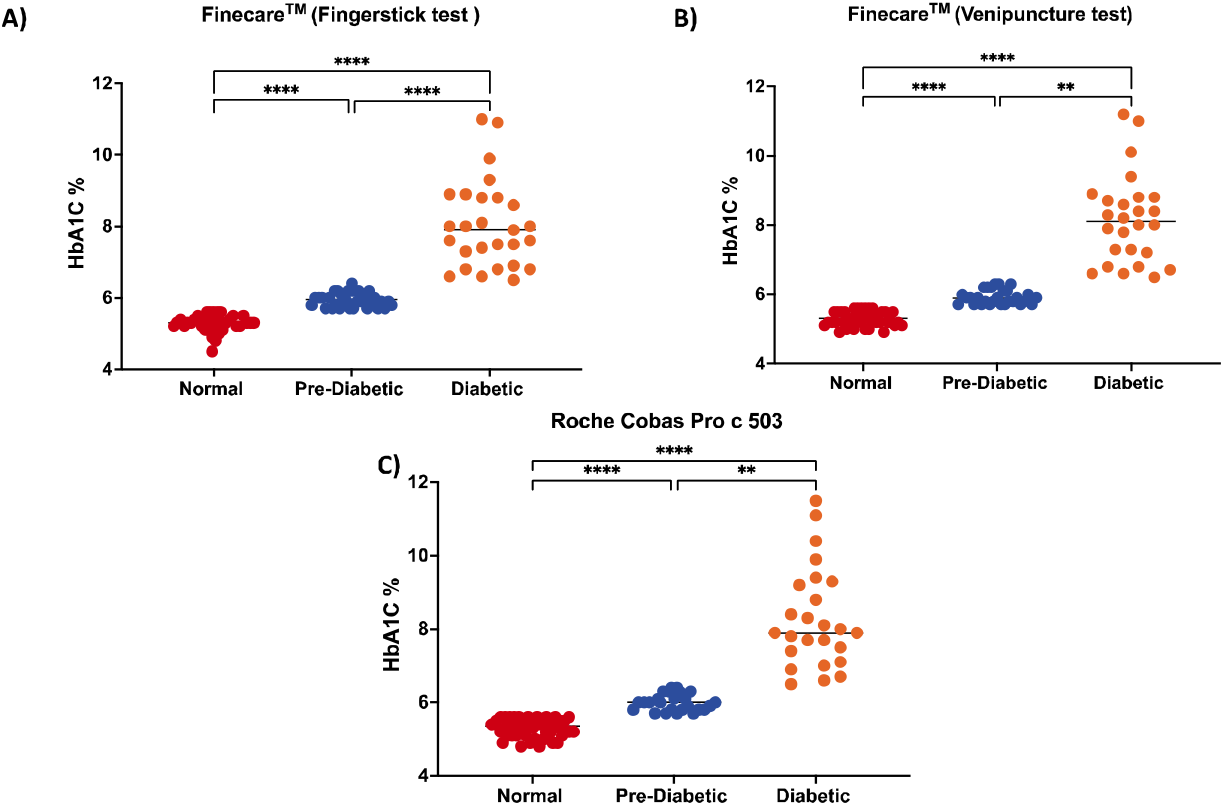
Distribution of numerical results obtained from fingerstick whole blood and venous blood using Finecare™ machine and the reference method; Roche Cobas Pro c 503. Results are represented as dot plots. Data are presented for 100 patients (normal <5.7%, pre-diabetic 5.7-6.4%, diabetic ≥6.5%) from each assay. Nonparametric Kruskal-Wallis test was used to compare the differences between groups.

### 3.6 Finecare™ showed excellent performance and reproducibility in another laboratory

To ensure the reproducibility of the results, the performance of Finecare™ was evaluated in another laboratory. Finecare™ showed 80% sensitivity and 100% specificity compared to the reference method (**Table 5**). Nevertheless, similar to our results, a very strong correlation was observed (*r*=0.97, *p*<0.0001) between Finecare™ and Roche Cobas Pro c 503 as shown in **Figure 5**.

**Table 5.** Comparison between Finecare™ (venepuncture) and Finecare™ (fingerstick) tests.

| Cut-off: 6.5% |  | REFERENCE METHOD<br>Roche HbA1c |  |  |
| --- | --- | --- | --- | --- |
|  |  | POSITIVE | NEGATIVE | TOTAL |
| Finecare™ | POSITIVE | 20 | 0 | 20 |
|  | NEGATIVE | 5 | 22 | 27 |
|  | TOTAL | 25 | 22 | 47 |

**Figure 5.**
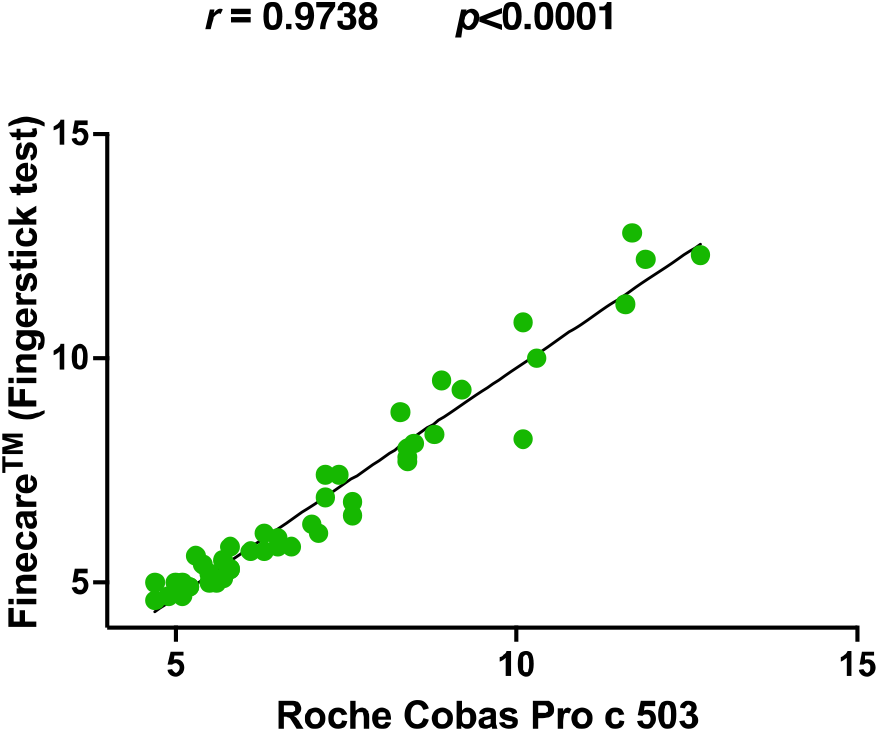
Pairwise correlation analysis and linear regression analysis of Finecare™ in comparison to Roche Cobas Pro c 503. Spearman correlation coefficient (r), and p value are indicated. Coefficient of determination (R^2^) was calculated to be 0.9496.

## 4. Discussion

This study validated the performance of Finecare™ HbA1c Rapid Quantitative Test used along with Finecare™ FIA System for quantitative determination of HbA1c in human blood. Our study is novel because there is no similar previous evaluation studies were found in the literature. The test is used as an aid to monitor long-term glycemic status in patients with diabetes mellitus. A total of 147 samples collected from two different labs (one private lab and another lab that belongs to the ministry of public health in Jordan) were used to evaluate the assays’ performance. To our knowledge, this is the first study conducted to validate the fluorescence-LFIA-based Finecare HbA1c test, which marks the novelty of this research work.

In this study, we demonstrated that Finecare™ results are comparable to the standard laboratory method Roche Cobas Pro c 503. No significant difference between Finecare™ and Roche Cobas Pro c 503 analytical analyser using the fingerstick or venous blood samples. The correlation and linear regression analyses between the readings obtained from Finecare™ (fingerstick and venous blood samples) and the reference method; Roche Cobas Pro c 503 was evaluated. Spearman correlation coefficients (r) demonstrated a statistically significant strong positive correlation between Finecare™ and Roche Cobas Pro c 503 with either fingerstick or venous blood samples (r>0.9, *p*<0.001). In addition, Cohen’s Kappa statistic denoted excellent agreement between Finecare™ and Roche Cobas Pro c 503. The excellent concordance between the POC Finecare™ and Roche Cobas Pro c 503 makes it an attractive alternative for the standard laboratory technique in non-laboratory setting.

One of the key advantages of Finecare™ is obtaining quantitative results within 5 minutes using fingerstick blood samples. Even though anti□HbA1c antibody are more stable and persistent in plasma samples, the fingerstick whole blood samples are more convenient and easier to use. Thus, we evaluated the efficacy of Finecare™ utilizing whole blood samples obtained through fingerstick to blood samples obtained via venipuncture. Using Finecare™, a very good correlation (*r*=0.905, *p*<0.0001) was detected between fingerstick and venous plasma samples. Our results validated the feasibility of utilizing whole blood sample from fingerstick for HbA1c detection and showed excellent concordance with venous blood samples values using Finecare™ test. Collection of fingerstick whole blood samples in Microtainer tubes is quick and easy. It eliminates the need for a phlebotomist, making it an attractive alternative for use in POC settings. The simplicity of performing LFIA with whole blood also eliminates the need for centrifugation and plasma separation steps, which reduce the cost and complexity of getting a quantitative result. The ability to rapidly, accurately, and affordably monitor HbA1c in diabetic patients is an important tool that will help in slowing the progression of type-2 diabetes particularly in patients residing in resource-limited areas.

In the present study, we were able to show that Finecare™ assay could be used efficiently for the long-term monitoring of HbA1c in diabetic patients. To ensure the reproducibility of the results, the performance of Finecare™ was evaluated in another laboratory. The assay performs with high reproducibility (80%) with the two runs repeats done on two different samples taken from two different laboratories. In addition, Finecare™ showed reproducible very high specificity (100%) compared to the reference method. Furthermore, a very strong correlation was also observed (*r*=0.97, *p*<0.0001) between Finecare™ and Roche Cobas Pro c 503. In conclusion, Finecare™ is a reliable assay and can be easily implemented for long-term monitoring of HbA1c in diabetic patients, particularly in none or small laboratory settings.

## Data Availability

All data produced in the present work are contained in the manuscript

## Author Contribution

Data analysis: GKN and NY. First draft writing: GKN and NY. Review and editing: M.M.G, A.M, and N.A.D

